# Impact of cardiac rehabilitation and treatment compliance after ST-segment elevation myocardial infarction (STEMI) in France, the STOP SCA+ study

**DOI:** 10.1101/2023.10.05.23296631

**Authors:** Emeline Laurent, Lucile Godillon, Marc-Florent Tassi, Pierre Marcollet, Stéphan Chassaing, Marie Decomis, Julien Bezin, Christophe Laure, Denis Angoulvant, Grégoire Range, Leslie Grammatico-Guillon

## Abstract

**Introduction:** Acute ST elevation myocardial infarction (STEMI) is one frequent and serious presentation of acute coronary syndrome. The STOP SCA+ study aimed to identify factors associated with negative outcomes one year after STEMI, particularly the impact of rehabilitation care and compliance.

**Methods:** Patients>18 years old hospitalized for STEMI management in five interventional cardiac centers in one French region (2.5 million inhabitants), between 2014 and 2018, were included. After a probabilistic matching with the National Health Insurance database (SNDS, 96% matching), compliance for cardiac tri-therapy was studied: aspirin, P2Y12 inhibitor statin. Factors associated with poor outcomes (ischemic complications, death) were analyzed using Cox modelling and those for the compliance by logistic regression.

**Results:** 3,768 patients were included, with 84% of primary percutaneous coronary intervention. At one year, 3,362 were prescribed a tri-therapy (89.2%) among whom 53% were compliant; 2,478 patients went to cardiac rehabilitation (65.8%). Death occurred in 130 patients and/or ischemic complication in 194 (total poor outcome 8.0%). Compliance was not associated with complications over the year (HR 1.16 [0.86-1.57]), while the absence of cardiac rehabilitation was (2.31 [1.73-3.08]). Additionally, cardiac rehabilitation was associated with compliance (OR 1.55 [1.34-1.79]).

**Discussion:** STEMI clinical evolution at one year is mainly favorable. Pejorative outcomes were scarce, and appeared to be related to patients’ characteristics, initial STEMI presentation, and no access to rehabilitation. Compliance part in patient health outcome will need further modelling to accurately study its impact. Matching clinical and medico-administrative databases proved to be relevant for assessing outcomes at a large scale.

**Key learning points:** *What is already known:*

- Although the compliance with a cardiac treatment and cardiac rehabilitation immediately after a myocardial infarction are key factors for improving the prognosis, less is known about compliance maintenance at one year.

*What this study adds:*

- At one year, few poor outcomes occurred and were not associated with compliance to the cardiac tri-therapy, while they were associated with the absence of cardiac rehabilitation.
- Matching two complementary clinical and medico-administrative databases proved to be reliable for assessing outcomes on a large scale (4,000 individuals over 5 years).

## Introduction

Cardiovascular diseases are a major cause of death in France, mainly due to coronary artery disease and its major complications, i.e. acute coronary syndrome [1–3]. Acute ST elevation myocardial infarction (STEMI) is a frequent and serious presentation, with a high risk of death in the acute phase but also a risk of later complications [4–7]. Currently, the reference treatment for STEMI consists in performing timely revascularization either by primary angioplasty (percutaneous intervention PCI) or by pharmacological fibrinolysis. PCI is the preferred alternative if it can be performed in an interventional cardiology centre (ICC) within less than 120 minutes between diagnosis and the interventional procedure [3, 8–10]. The earlier the procedure is performed, the shorter the myocardial ischemia stands and the better the functional recovery and the survival rate [6, 9, 11, 12]. The prescription and compliance with a secondary prevention cardiac treatment after a STEMI are also key factors for improving the prognosis. Indeed, drug management after myocardial infarction (BASI: Beta-blocker, Anti-platelet therapy, Statin, Angiotensin-converting enzyme Inhibitor) must be optimized and maintained over time to prevent ischaemic recurrences and death, with a high-level recommendation for a dual antiplatelet therapy DAPT (including aspirin and P2Y12 inhibitor) in the following one-year period after PCI, along with a long-term LDL-cholesterol lowering therapy [13, 14].

Using an automated procedure to monitor the immediate outcomes of STEMI patients and their follow-up, including medication intake, could provide high-quality data without significant cost and time waste, as previously demonstrated [15]. Therefore, in the French region of Centre-Val de Loire (which has approximately 2.5 million inhabitants and 6 ICCs), an electronic registry was established in 2014 (CRAC registry). This registry is fully integrated with the coronary activity report software and has an automatic daily update. By the end of 2018, it had included a total of 4,204 patients [3, 8]. Its national extension, France PCI, is accelerating because it represents a source for real-life studies to increase current scientific knowledge at low cost (www.francepci.com).

The CRAC registry has already provided insights into several factors that impact STEMI mortality, such as patient age, high Killip score at admission, or the positive impact of calling the French medical emergency number [3, 9]. However, some registry variables related to the one-year follow-up, particularly therapeutic compliance, are collected by phone calls, which can sometimes be incomplete (not collected for 13% of the patients due to death or lost to follow-up). Therefore, to improve the quality and completeness of the data and minimize loss to follow-up, matching the clinical CRAC registry data with the National Health Insurance data (*Système National des Données de Santé*, SNDS) would allow for a comprehensive approach to compliance and one-year outcomes for STEMI patients.

The aim of the cohort study, STOP-SCA+, was to describe compliance to secondary prevention cardiac tri-therapy (i.e. aspirin, P2Y12 inhibitor, and statin) of patients over the one-year period after STEMI, by matching the CRAC registry with the SNDS database. The secondary objectives were to identify i) the factors associated with poor outcomes (death or ischemic complications) at one year, especially the impact of rehabilitation care and cardiac tri-therapy compliance; and ii) the factors associated with compliance to the secondary prevention cardiac tri-therapy.

## Methods

This study was a retrospective, multicentre and observational cohort study with a probabilistic matching (in the absence of a common direct identifier for each patient) between two databases, the CRAC registry and the medico-administrative SNDS database.

Eligible patients were all patients aged over 18 years old, who underwent a coronary angiography or PCI at the acute phase of a STEMI, in five French ICC, between January 1^st^ 2014 and December 31^st^ 2018, as identified from the CRAC registry. Exclusion criteria were the absence of consent, death occurring during the inclusion ICC hospital stay, or the absence of matching between CRAC registry and SNDS data.

### Data sources and matching

The CRAC Registry includes up to 150 variables for each patient, including pre-admission, hospital and follow-up data, as described elsewhere [3]. The SNDS includes the vital status and all reimbursement data for out-of-hospital drug prescriptions and consultations, along with all hospitalisations in public and private sectors in France (although without detail about drug consumptions), linked by a unique encrypted patient number [15, 16]. The probabilistic matching between the CRAC registry and the SNDS was performed as described in the Supplementary material S1 (Tables S1.1 and S1.2). Each of the seven matching steps had excellent performance parameters, with predictive positive values reaching >99.9% for each step (the lowest parameter performance was a sensitivity estimation at 82% for one step involving only a few cases). Overall, 96% of the CRAC registry cases could be matched with their counterpart SNDS unique cases (Supplementary figure S1.3).

This study was compliant with French regulation about data protection and authorized by the French Data Protection Board (*Commission Nationale de l’Informatique et des Libertés*, CNIL), decision DR-2021-025 (January 28th, 2021). This study required neither information nor consent of the included individuals.

### Main outcome and variables of interest

The main outcome was defined as the occurrence of an ischaemic complication or death during the first year after being discharged alive from the ICC. The ischaemic complications were extracted from the CRAC registry: stent thrombosis, myocardial infarction, non-haemorrhagic stroke, unplanned revascularisation, target lesion revascularisation. The vital status was checked using both CRAC registry and SNDS data up to one year after discharge. For compliance assessment, three therapeutic classes were considered: aspirin, P2Y12 inhibitors, and statin. These medications were considered separately or combined as (i) DAPT (aspirin + P2Y12 inhibitor), or (ii) tri-therapy: DAPT + statin. Patients with one anti platelet agent (aspirin or P2Y12 inhibitor) associated with an anticoagulant were assimilated to DAPT patients. This association allowed to consider patients with a contraindication to aspirin. For each medication, the SNDS database provides the dates of delivery, which was used as a proxy for the date of administration. For cardiac rehabilitation, all hospitalisations with at least one diagnosis code Z50.0 as main care were considered, including outpatient stays.

The other variables of interest included socio-demographics characteristics (age, sex, social deprivation according to a deprivation score “Fdep” for each French area [17], divided in 5 quintiles), underlying conditions (such as hypertension, diabetes mellitus), medical history (such as myocardial infraction, stroke), clinical presentation at onset (including the Killip score), procedural data, left ventricular ejection fraction (LVEF) at discharge and haemorrhagic complications over the year after discharge; extracted from the CRAC registry. Other cardiac drugs delivered (beta-blockers, angiotensin-converting enzyme inhibitor) and general practitioner (GP) consultations were extracted from the SNDS over the year after discharge. Rehospitalizations were studied and defined as acute care hospitalisations with at least one night, occurring after discharge from the index stay.

### Statistical analyses

First, patients, clinical presentations and one-year outcomes were described in terms of numbers and percentages/frequencies. The median time intervals between the discharge from STEMI hospital stay and the admission in a rehabilitation center were calculated, along with their first (Q1) and third (Q3) quartiles.

Second, the factors associated with the occurrence of an ischaemic complication or death were identified, with i) log-rank tests in bivariate analyses, then ii) Cox modelling in multivariate analysis, including cardiac rehabilitation and tri-therapy as time-dependent variables. For time-dependent tri-therapy, a period of exposure was defined as a period covered by a delivery for all three medications, whereas a non-exposure period corresponded to a period with at least one medication missing, out of three. Each hospitalisation period, both in acute or rehabilitation care, was considered as a full-exposure period for deliveries registered before or after the hospitalisation, as the delivery is provided by the healthcare facility and not reported in the SNDS. For the final model (see below), the absence of interaction between cardiac rehabilitation and compliance was checked. Hazard-ratios (HR), along with their 95% confidence interval (_95%_CI) were reported.

Third, after checking the absence of interaction between cardiac rehabilitation and compliance in the previous model, a secondary analysis was performed to assess the factors associated with the compliance (proportion of days covered PDC ≥80%) for cardiac tri-therapy, in a subgroup of patients without major adverse cardiac or cerebral event during the subsequent year. The PDC was defined as the proportion of days covered by drug deliveries up to one year (or up to death before one year). For this analysis, a good compliance was defined by a PDC ≥80% [18–20]. After a bivariate analysis using Chi-square tests, a multivariate logistic regression model was performed, giving odds-ratio (OR), along with their _95%_CIs.

For all multivariate analyses, all variables with p<0.2 were included in the initial model, the final model being selected through a descending stepwise process, to select variables with p<0.05, along with clinically pertinent variables, sex and age. For all tests, the threshold for statistical significance was set to 5%. All analyses were conducted using SAS® Enterprise Guide software (SAS® Institute Inc., Cary, NC, USA), version available on the national SNDS portal at the time of the analyses.

## Results

From the 4,179 eligible patients from the CRAC registry, 3,768 were eventually included (90% of the eligible patients): discharged alive with available SNDS data over the follow-up year (Supplementary S1 - figure S1.3).

The mean age was 62 years old [min-max 18-96], with three times more men than women (table 1). Over half of the patients were smokers (53%), 40% had a high blood pressure, 22% had a family history of coronary disease. Obesity was observed in 21% of the cases and diabetes mellitus in 14% of the cases. Among the cohort patients, 93% had a revascularization (84% had a primary angioplasty). At discharge, 20% of the patients had a LVEF <40% (table 1).

**Table 1.** Patients’ characteristics and hospital management – The STOP-SCA+ study.

|  | Patients included |  |  |
| --- | --- | --- | --- |
|  | n | % | missing |
| **Total** | **3,768** | **100** |  |
| **Age, years (mean [min-max])** | 62 [18-96] |  | 0 |
| **sex-ratio M/F** | 3.0 |  | 0 |
| **Comorbidities** |  |  |  |
| Overweight ($25 \leq \text{BMI} < 30\text{kg/m}^2$) | 1,599 | 42.5 | 4 |
| Obesity ($\text{BMI} \geq 30\text{kg/m}^2$) | 789 | 21.0 | 4 |
| Renal impairment | 52 | 1.4 | 36 |
| High blood pressure | 1,510 | 40.3 | 25 |
| Diabetes | 519 | 13.9 | 25 |
| Smoking (current or past) | 2,006 | 53.5 | 18 |
| **Medical history** |  |  |  |
| TCA/myocardial infarction/coronary bypass | 465 | 12.3 | 0 |
| Stroke | 94 | 2.5 | 3 |
| Peripheral vascular disease | 99 | 2.6 | 10 |
| Family history of coronary disease | 797 | 21.8 | 115 |
| **Killip = 3 or 4 at the admission** | 132 | 3.6 | 72 |
| **Reperfusion procedure** | 3,497 | 92.8 | 0 |
| Primary angioplasty | 3,171 | 84.2 |  |
| Isolated fibrinolysis | 54 | 1.4 |  |
| Secondary angioplasty | 272 | 7.2 |  |
| Isolated coronary angiography | 271 | 7.2 |  |
| **LVEF at discharge < 40%** | 665 | 19.5 | 362 |

BMI: Body Mass Index; LVEF: Left Ventricular Ejection Fraction; TCA: Transluminal Coronary Angioplasty

During the one-year follow-up, 98% of the patients were prescribed at least once aspirin, 90% a P2Y12 inhibitor and 97% a statin, resulting in 89% of the patients with at least one delivery of a cardiac tri-therapy (n=3,362). Among these patients, 53% were compliant to their tri-therapy (PDC ≥80%) (figure 1, S2 and S3).

**Figure 1.**
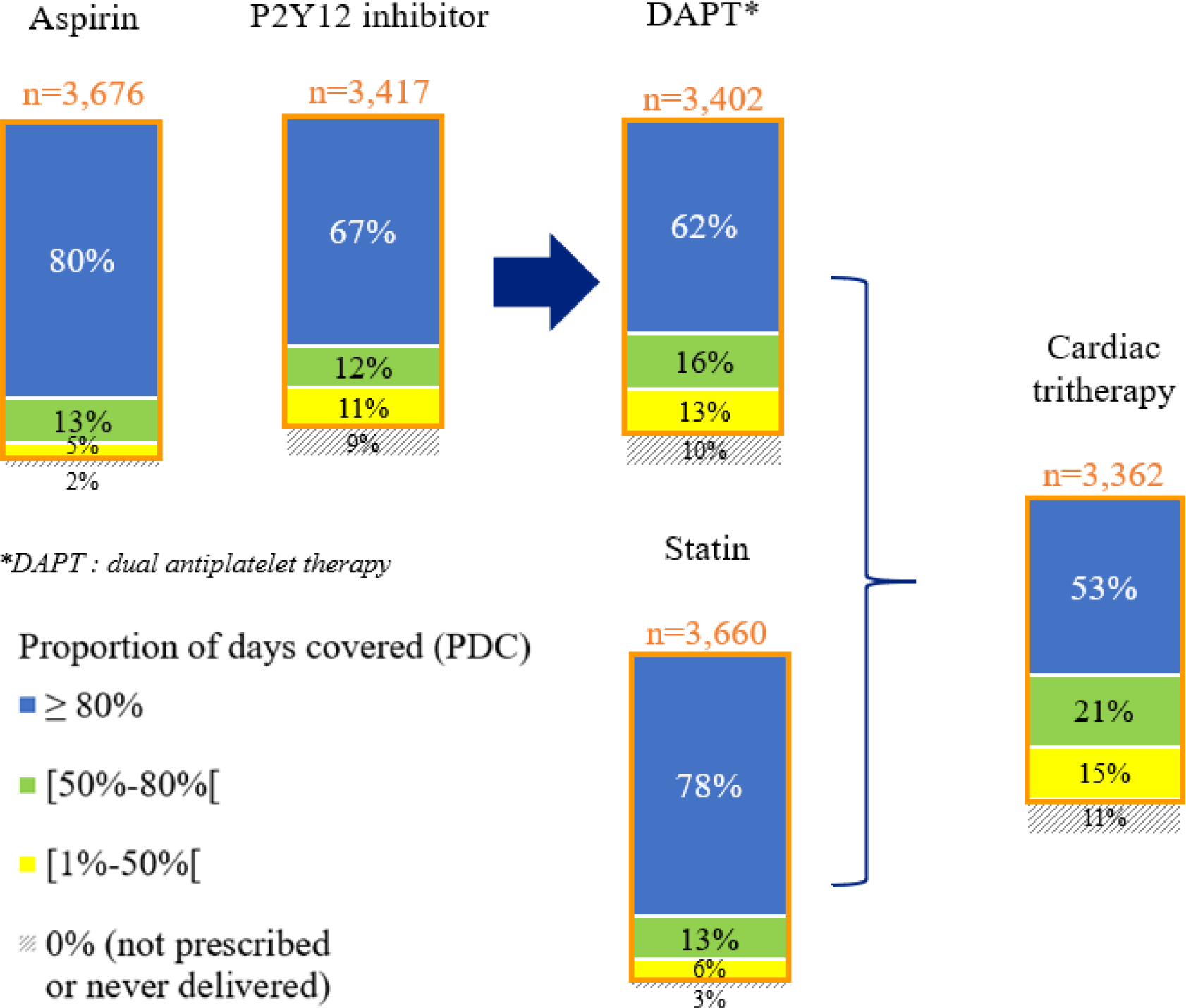
Cardiac drug compliance at one year after STEMI – the STOP-SCA+ study. *PDC: proportion of days covered by the studied medication*

At one year, 66% of the STEMI patients had access to a cardiac rehabilitation centre (table 2). The median time interval between the STEMI patient discharge from acute care setting to admission to a cardiac rehabilitation center was 8 days (Q1: 1 day - Q3: 20 days). The median time interval between the reperfusion and the admission in a cardiac rehabilitation center was 14 days (Q1: 8 days-Q3: 26 days).

**Table 2.** Patients’ outcome at one year after STEMI – The STOP-SCA+ study.

| The STOP-SCA+ study: 1-year outcome | Patients included |  |
| --- | --- | --- |
|  | n | % |
| **Total** | **3,768** | **100** |
| **One-year re-hospitalisations** |  |  |
| Acute care (full hospitalisation) | 1,817 | 48.2 |
| with vascular disease | 1,327 | 35.2 |
| with emergency admission for vascular disease | 299 | 7.9 |
| Rehabilitation care |  |  |
| In-hospital stay | 1,787 | 47.4 |
| Outpatient stay | 994 | 26.4 |
| with cardiac rehabilitation (in or outpatient stay)* | 2,478 | 65.8 |
| **Ischaemic complication\*\* or death** | 303 | 8.0 |
| Ischaemic complication | 194 | 5.1 |
| Death | 130 | 3.5 |
| **Haemorrhagic complication** | 73 | 1.9 |

\* ICD-10 diagnosis code Z50.0 "Cardiac rehabilitation" as the main care registered
\*\* stent thrombosis, myocardial infarction, non-haemorrhagic stroke, unplanned revascularisation, target lesion revascularisation

Over the one-year follow-up, 303 patients (8.0%) had a pejorative outcome: ischaemic complication (n=194; 5.1%) and/or death (n=130; 3.5%) (table 2). This proportion varied according to the patients’ characteristics and management (table 3).

**Table 3.** Ischaemic complication and/or death at one year after STEMI according to their characteristics and management – The STOP-SCA+ study.

|  | Total | Among whom,<br>ischaemic<br>complication<br>and/or death<br>(n=303) | bivariate<br>p-value<br>(log-rank) |
| --- | --- | --- | --- |
|  | n | % (in line) |  |
| **All patients** | 3 768 | 8,0 |  |
| **Age ≥65 years old** | 1 601 | 10,7 | <0.01 |
| **Women** | 936 | 7,7 | 0,67 |
| **Comorbidities** |  |  |  |
| Obesity (BMI ≥ 30kg/m2) | 789 | 7,2 | 0,34 |
| Renal impairment | 52 | 26,9 | <0.01 |
| High blood pressure | 1 510 | 10,1 | <0.01 |
| Diabetes | 519 | 14,6 | <0.01 |
| Smoking (current or past) | 2 006 | 7,7 | 0,33 |
| **Medical history** |  |  |  |
| TCA/myocardial infarction/coronary bypass | 465 | 12,9 | <0.01 |
| Stroke | 94 | 13,8 | 0,03 |
| Peripheral vascular disease | 99 | 19,2 | <0.01 |
| Family history of coronary disease | 797 | 7,5 | 0,56 |
| **Killip = 3 or 4** | 132 | 17,4 | <0.01 |
| **Reperfusion procedure** |  |  | 0,83 |
| Primary angioplasty | 3 171 | 8,1 |  |
| Isolated fibrinolysis | 54 | 11,1 |  |
| Secondary angioplasty | 272 | 7,7 |  |
| Isolated coronary angiography | 271 | 7,4 |  |
| **LVEF at discharge <40%\*** | 665 | 12,2 | <0.01 |
| **No cardiac rehabilitation** | 1 290 | 12,3 | <0.01 |

\* missing data: n=362
BMI: Body Mass Index; LVEF: Left Ventricular Ejection Fraction; MI: Myocardial Infarction; TCA: Transluminal Coronary Angioplasty

In multivariate analyses, the significant factors associated with an ischaemic complication and/or death were being a female, renal impairment, a pejorative Killip, a LVEF <40% at discharge and the absence of cardiac rehabilitation (figure 2, S4). On the contrary, the compliance was not significantly associated with the occurrence of a pejorative outcome. No interaction was found between cardiac rehabilitation and cardiac tri-therapy compliance. The social deprivation estimated through the Fdep index was not associated with pejorative outcomes in bivariate analysis (results not reported), thus not included in the multivariate analysis. Studying the factors associated with cardiac tri-therapy compliance (PDC ≥80%), cardiac rehabilitation was a contributive factor (OR=1.55 [1.34-1.79]) (figure 3, S5), along with a younger age, being a male, the absence of medical history of stroke or MI (figure 3).

**Figure 2.**
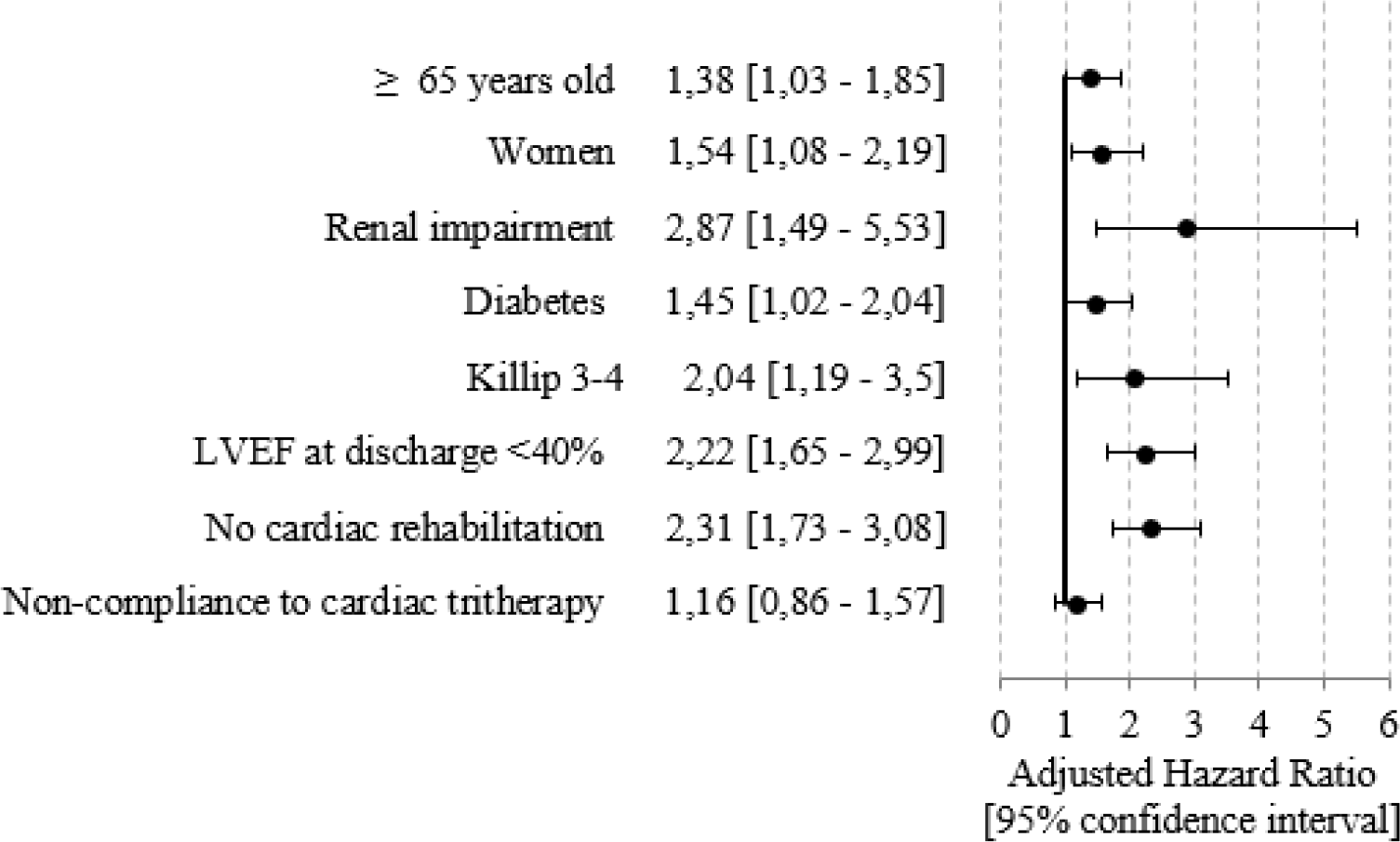
Factors associated with an ischaemic complication and/or death at one year after STEMI – the STOP-SCA+ study. *LVEF: Left Ventricular Ejection Fraction*

**Figure 3.**
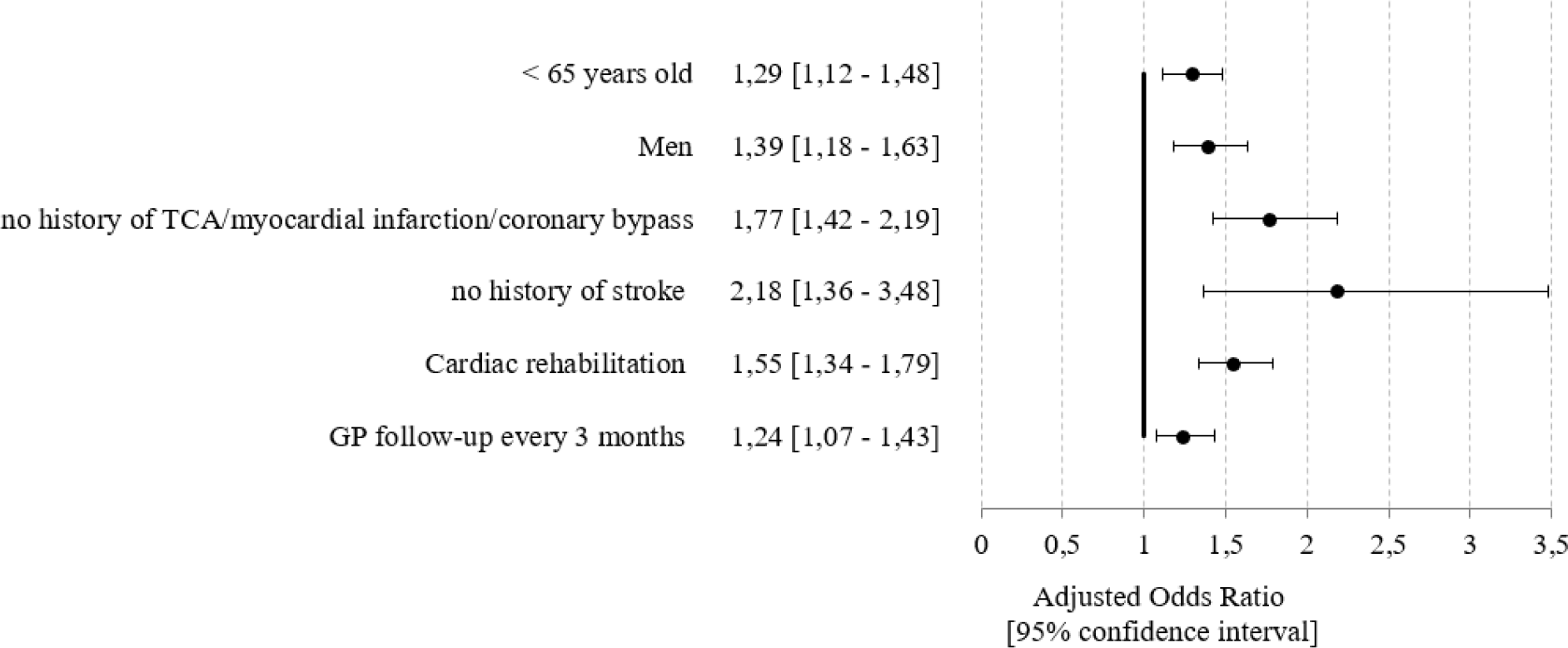
Factors associated with compliance for the cardiac tri-therapy (PDC ≥80%) at one year after STEMI – the STOP-SCA+ study *GP: General Practicioner; TCA: Transluminal Coronary Angioplasty*

## Discussion

This study showed that the overall outcome at one year after a STEMI is satisfying, with few complications nor death. Only 8% of the patients had poor outcome, mainly related to their pre-existing conditions, the initial severity of the STEMI, but also associated with the absence of cardiac rehabilitation, whereas the observed compliance to the cardiac tri-therapy wasn’t significantly associated. Here, we found that almost all the STEMI patients were prescribed at least once aspirin (98% of patients), a P2Y12 inhibitor (90%) or a statin (97%), resulting in 89% with at least one tri-therapy prescription, but only one half of them had a PDC≥80. We also highlighted the interest to consider the medical compliance as both a static parameter, such as the PDC over a defined period, and a dynamic one, such as a time-dependent variable, whatever the total duration of deliveries. Our data are in line with the FAST-MI registry results that showed a significant risk reduction of one-year mortality in a French cohort of patients undergoing cardiac rehabilitation after admission for acute myocardial infarction [21, 22].

The originality and interest of our study are multiple, as regards these assessments. First, our result was the first to date matching two complementary databases: the clinico-biological data from the CRAC registry (subgroup of France-PCI), including a large STEMI population of nearly 4,000 individuals over 5 years, and the medico-administrative data of the SNDS, allowing to increase the completeness and accuracy of the results. The SNDS proved its epidemiological interest, allowing to catch complete and large-scale data, especially after discharge in outpatient care, which usually require an onsite follow-up led by local research technicians, impacting cost and resulting in an underestimation of the results due to lost-of follow-up / dead patients. The matching was reliable and efficient and could provide an estimation of the operating needed resources assessed in real-life settings. However, some limitations were highlighted as regards the definition of the compliance through this medico-administrative database [19, 23]. The SNDS contains the dispensations and not the administration, and delivery discontinuations cannot be identified, resulting in a possible misestimation of compliance; under-estimation for patients for whom the delivery was stopped due to health or tolerance issues, vs. self-discontinuation by patients who do not feel the usefulness of the treatment, in the absence of symptoms, i.e., overestimation of compliance if delivered but not taken. Moreover, the SNDS medico-administrative database did not allow to take into account a number of other confounders that could affect both medication compliance and outcomes, such as environmental factors. However, social deprivation was considered through the Fdep index.

Our results also provided some insight where no clear consensus can be found as regards the duration of DAPT or statin after a STEMI: according to the 2017 ESC guidelines, “multiple studies have shown that shortening DAPT to 6 months, compared with 12 months or longer, reduces the risk of major bleeding complications, with no apparent trade-off in ischaemic events”, whereas “two major studies have shown the benefit towards reduction of non-fatal ischaemic events in patients receiving longer than 12 months of DAPT”, thus concluding that “no formal recommendations are possible for the use of clopidogrel or prasugrel beyond 1 year.”[10, 13] As regards the long-term prescription of statin, although highly recommended, no duration is indicated in the ESC guidelines other than “lipids should be re-evaluated 4–6 weeks after the [acute coronary syndrome] to determine whether the target levels have been reached and regarding safety issues; the lipid lowering therapy can then be adjusted accordingly.” Long term follow-up of patients enrolled in clinical trials testing lipid lowering therapy in high-risk patients advocates for a beneficial risk ratio to benefit of prolonged LDL lowering therapy [24] Other post-STEMI recommended treatments included in the BASI [13, 14], were not considered for this study, due to numerous posology adaptations over time, not allowing to reliably assess compliance.

Second, our study took into account post-STEMI cardiac rehabilitation through the SNDS database, which appeared to be a major factor to improve the outcome at one year, as previously suggested, although a strong causal model remains to be built [25, 26]. Our results were particularly encouraging, as two-thirds of the patients benefited from cardiac rehabilitation, which was higher than previous data, where about a third of coronary patients in Europe receives any form of cardiac rehabilitation and only a 25% in the USA [20, 21]. In France, Puymirat *et al* demonstrated the benefits of cardiac rehabilitation after admission for acute myocardial infarction with a risk reduction of one-year mortality [21].

The CRAC registry has also some limitations: the inclusion criterion is limited to patients admitted in ICC. Thus, STEMI patients without any invasive procedure, even if rare, are not analysed, conducing to a selection bias. However, previous studies estimated that hospitalized STEMI patients without coronary angiography corresponded to less than 5% in France [20, 27]. This regional registry including 5 ICC is now extended to over 40 ICC across the country in France PCI as the SNDS is part of the French Health data hub (www.francepci.fr), which will allow to extend our findings to a national level in a close era.

## Conclusions

This study showed that patients admitted for emergency revascularization at the acute phase of a STEMI mainly had favourable outcomes at one year, with few complications or fatality. Pejorative outcomes appeared to be related to patients’ characteristics, initial STEMI presentation, and access to rehabilitation care, whereas no impact could be observed for non-compliance. Matching two complementary clinical and medico-administrative databases proved to be reliable for assessing outcomes on a large scale. This permanent and multicentric database with systematic long-term follow-up in IC provides excellent quality of data and immediate feedbacks at a large scale, without high cost or changing usual practice due to one full integrated electronic medical report. This large reliable database could soon provide care quality improvement linked to benchmarking and promote scientific robust studies.

## Data Availability

The data underlying this article cannot be shared publicly due to privacy and legal reasons, according to the decision from the French Data Protection Board (Commission Nationale de l'Informatique et des Libertés, CNIL), decision DR-2021-025 (January 28th, 2021).

## Data Availability Statements

The data underlying this article cannot be shared publicly due to privacy and legal reasons, according to the decision from the French Data Protection Board (*Commission Nationale de l’Informatique et des Libertés*, CNIL), decision DR-2021-025 (January 28th, 2021).

